# Prehospital activation of the cardiac catheterisation laboratory in ST-segment elevation myocardial infarction (STEMI) for primary percutaneous coronary intervention (PCI)

**DOI:** 10.1101/2023.05.16.23290073

**Authors:** Michael L Savage, Karen Hay, William Volbon, Tan Doan, Dale Murdoch J, Christopher Hammett, Rohan Poulter, Darren L Walters, Russell Denman, Isuru Ranasinghe, Owen Christopher Raffel

## Abstract

**Background:** Prehospital activation of the cardiac catheter laboratory is associated with significant improvements in ST-segment elevation myocardial infarction (STEMI) performance measures. However, there is equivocal data, particularly within Australia regarding its influence on mortality. We assessed the association of prehospital activation on performance measures and mortality in STEMI patients treated with primary percutaneous coronary intervention (PCI) from the Queensland Cardiac Outcomes Registry (QCOR).

**Methods:** Consecutive ambulance transported STEMI patients treated with primary PCI were analysed from 1^st^ January 2017 to 31^st^ December 2020 from the QCOR. The total and direct effects of prehospital activation on the primary outcomes (30-day and 1-year cardiovascular mortality) were estimated using logistic regression analyses. Secondary outcomes were STEMI performance measures.

**Results:** Among 2498 patients (mean age: 62.2 ± 12.4 years; 79.2% male), 73% underwent prehospital activation. Median door-to-balloon (DTB) time (34mins [26-46] vs 86 mins [68-113]; p<0.001), first-electrocardiograph-to-balloon (ECGTB) time (83.5 mins [72-98] vs 109 mins [81-139]; p<0.001), and proportion of patients meeting STEMI targets (DTB<60mins 90% vs 16%; p<0.001), ECGTB<90mins (62% vs 33%; p<0.001) were significantly improved with prehospital activation. Prehospital activation was associated with significantly lower 30-day (1.6% vs 6.6%; p<0.001) and 1-year cardiovascular mortality (2.9% vs 9.5%; p<0.001). After adjustment, no prehospital activation was strongly associated with increased 30-day (OR: 3.6; 95%CI: 2.2-6.0, p<0.001) and 1-year cardiovascular mortality (OR: 3.0 (95%CI:2.0-4.6; p<0.001).

**Conclusions:** Prehospital activation of cardiac catheterisation laboratory for primary PCI was associated with significantly shorter time to reperfusion, achievement of STEMI performance measures and lower 30-day and 1-year cardiovascular mortality.

**Clinical Perspective:**

- In patients who suffer STEMI, prehospital activation of the cardiac catheter laboratory and initiation of medical therapy is associated with shorter time to reperfusion, greater achievement of performance measures and lower cardiovascular mortality
- This study adds to the existing literature and demonstrates that a standardised prehospital activation strategy can be implemented on a large scale
- Widespread implementation of standardised prehospital activation strategies may offer opportunity to expedite STEMI care and improve outcomes

## Introduction

Cardiovascular mortality remains one of the leading causes of death worldwide, with majority of deaths occurring following acute myocardial infarctions [1]. Early identification and treatment of acute ST-segment elevation myocardial infarction (STEMI) is crucial with the primary aim is to minimise the delay to reperfusion of the culprit coronary artery. The preferred method of reperfusion, if it can be achieved within 120mins of diagnosis, is primary percutaneous coronary intervention (PCI). [1]. Historically, reducing the in-hospital delay to treatment or door-to-balloon (DTB) time has been the primary focus of improving outcomes in with primary PCI, and is associated with lower short- and long-term mortality [2–4]. Several hospital-based strategies have seen significant reductions in median DTB times since their implementation [5]. However, despite these reductions in DTB, overall mortality following STEMI has not significantly improved over time with primary PCI [2, 4].

There has been a more recent shift to improve patient outcomes following STEMI by focusing on the collaboration with prehospital emergency medical services and incorporating strategies to reduce the prehospital delay to treatment. Implementation of several prehospital strategies both locally and internationally such as the Pre-Act initiative have demonstrated reductions in prehospital delays, further reductions in DTB time and total ischaemic time, and reduced false positive catheterisation laboratory (CCL) activations when protocol driven prehospital STEMI pathways are implemented [6–11]. The impact of prehospital notification of STEMI and prehospital activation of the CCL on mortality however remains mixed with either lower [7, 8, 12-15] or equivalent [16, 17] outcomes reported both across Australia and internationally. The Australian Acute Coronary Syndrome guidelines recommend primary PCI as the preferred reperfusion strategy if it can be performed within 90 minutes of first medical contact (FMC) [18] yet do not specify prehospital notification of STEMI by paramedics and/or prehospital activation of the CCL as strategies for achieving performance targets.

Significant variability exists in the design, implementation and utilisation of prehospital strategies particularly within Australia [11, 14, 15]. Prehospital emergency care within Queensland, Australia is provided by the Queensland Ambulance Service (QAS), which is a single, state-wide, government-funded service. The state-wide prehospital direct primary PCI referral pathway and prehospital activation strategy has been previously described in detail [14, 19] (Supplement 1). The aim of this study was to examine the impact of a well-established state-wide prehospital activation strategy on achievement of STEMI performance measures and mortality in patients presenting with STEMI who received primary.

## Methods

This multicentre study consisted of consecutive STEMI patients over a 4-year period (1^st^ January 2017 – 31^st^ December 2020) who were transported by ambulance to one of eight participating primary PCI facilities in Queensland (Supplement 2) and underwent primary PCI. Queensland, a state within north-eastern Australia, covers an area of over 1.8 million square kilometers (695 thousand square miles) and services a population of approximately 5.3 million people. Patients who experienced an in-hospital STEMI, underwent inter-hospital transfers or self-presented were excluded from analysis. Patients with out-of-hospital cardiac arrest (OHCA) and emergency pre-procedural intubation with unknown neurological status were also excluded (Figure 1). Comparisons were performed between ambulance transported STEMI patients who underwent the direct primary PCI referral and prehospital CCL activation pathway with ambulance transported STEMI patients who did not have direct primary PCI referral and prehospital activation of the CCL.

**Figure 1.**
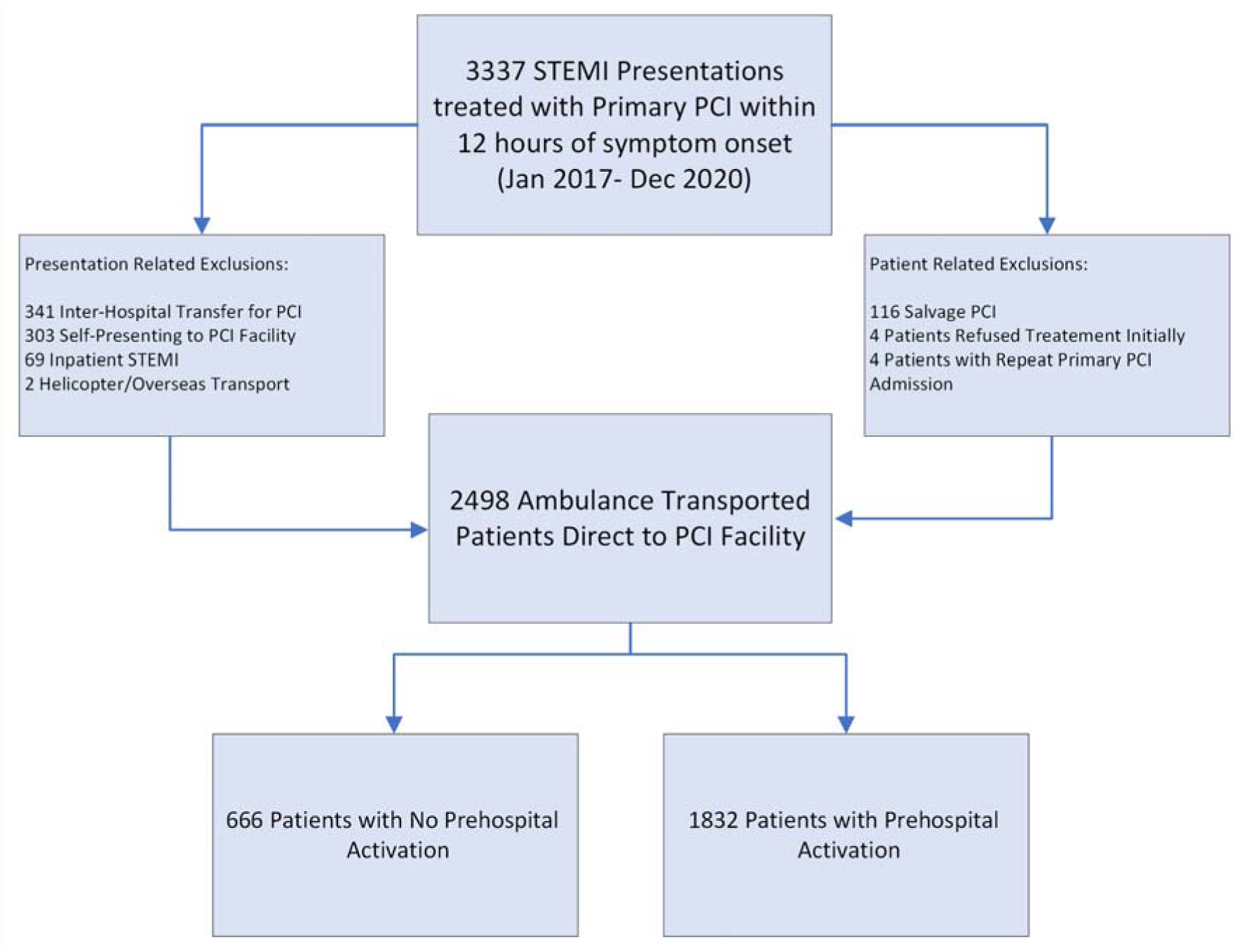
Study inclusion flow diagram. PCI – Percutaneous Coronary Intervention, STEMI – ST-segment elevation myocardial infarction, Salvage PCI – Out of hospital cardiac arrest with emergency intubation.

The state-wide prehospital activation strategy involves STEMI diagnosis, PCI referral subject to reperfusion checklist (Supplement 1)and initiation of medical therapy by paramedics upon direct telephone consultation with the on-call interventional cardiologist, who activates the CCL team prior to hospital arrival. Patient demographic characteristics, procedural variables, STEMI performance measures and clinical endpoints of 30-day mortality and 1-year mortality were compared between groups. Patients in this study who underwent prehospital activation pathway were administered prehospital concomitant antiplatelet therapy (aspirin 300mg orally in combination with ticagrelor 180mg orally or clopidogrel 600mg orally) and anticoagulant therapy (heparin 5,000 units intravenously) unless contraindicated, by the paramedics immediately post PCI referral. Whereas those who did not undergo prehospital activation, did not have administration of heparin nor ticagrelor/clopidogrel until review in the emergency department.

## Data Sources

The Queensland Cardiac Outcomes Registry (QCOR) is a government funded central, state-wide data registry of which all public hospital cardiac catheter laboratories contribute patient information and procedural data. There are 8 public hospitals who participate, 3 metropolitan centres and 5 regional centres (Supplement 2). Patient demographics and periprocedural data were obtained from the QCOR and hospital electronic medical record, with data linkage from the Queensland Hospital Admitted Patient Data Collection from the Queensland Health Statistical Services Branch. Prehospital data were obtained from data linkage with the QAS STEMI database. Mortality and cause of death data were obtained utilising data linkage with the Queensland Registry of Births Deaths and Marriages. This study was approved by the Human Research Ethics Committee of the Prince Charles Hospital (LNR/2020/TPCH/75136) with a waiver of consent obtained. Because of the sensitive nature of the data collected for this study, requests to access the dataset from qualified researchers trained in human subject confidentiality protocols may be sent to the relevant data custodians within Queensland Health.

## Data definitions

DTB time was defined as the time in minutes from arrival at the PCI facility to the use of the first wire, balloon or aspiration thrombectomy device in the PCI procedure. First Medical Contact (FMC) for all patients was defined as initial paramedic contact. First Diagnostic Electrocardiograph (FDECG) was defined as the first electrocardiograph which demonstrated ST-segment changes consistent with the diagnosis of STEMI. Out of hours (OH) presentation was defined as between 1700-0800 hours during weekdays and during the weekend. Cardiovascular mortality was defined as any deaths resulting from the combination of ischaemic heart disease, cardiac failure, cerebrovascular disease, peripheral artery disease and aortic aneurysm.

## Statistical Analysis

Continuous variables were compared between groups using an independent t-test or Wilcoxon’s rank-sum test as appropriate and categorical variables were compared using Pearson’s chi-square test. Continuous variables were categorised using clinically meaningful cut-points. A causal diagram (Supplement 3) was constructed *a priori* to depict postulated inter-relationships between variables of interest, confounders and mortality. The Dagitty [20] online user interface was used to select appropriate adjustment sets of covariates to estimate the total and direct effects of prehospital activation on 1-year mortality [21]. Confounders of interest in the relationship between prehospital activation and mortality comprised patient characteristics (age, sex) and presentation characteristics (cardiac arrest pre-PCI, regional hospital, out of hours presentation). Total effect estimates for the effect of prehospital activation on 30-day and 1-year mortality were derived from multivariable logistic regression models adjusted for confounders in the minimally sufficient adjustment set (age, sex, cardiac arrest pre-PCI, regional hospital, out of hours presentation) while direct effect estimates were additionally adjusted for the mediator (FDECG to balloon). Survival over time was explored graphically using Kaplan Meier methods. Analyses were performed using the Stata statistical software package (StataCorp. 2017. *Stata Statistical Software: Release 15*. College Station, TX: StataCorp LLC).

## Results

### Cohort Characteristics

A total of 2,498 patients were included, of which 1,832 (73.3%) patients underwent in the prehospital activation and 666 (26.7%) who did not have prehospital activation. The mean age of the patients was 62.2 years (SD:12.4) and 1978 (79.2%) were male (Table 1). The proportion of STEMI presentations receiving prehospital activation did not significantly differ over the course of the study period (p=0.072). Patients with prehospital activation were on more likely to be male, have dyslipidaemia and less likely to have a cardiac arrest pre-PCI (Table 1). The mean age, use of radial arterial access and drug eluting stents (DES) were similar between groups and did not significantly change over the study period. Rates of Intra-aortic balloon pump and temporary pacing wire requirements were low and similar between groups. Prehospital activation was less likely to occur out of hours compared to in hours (IH) (OH 70% vs IH 78%; p<0.001) and in regional centres (Metropolitan 80% vs Regional 65%, p<0.001). Regional and metropolitan centres are listed in supplement 2.

**Table 1:**
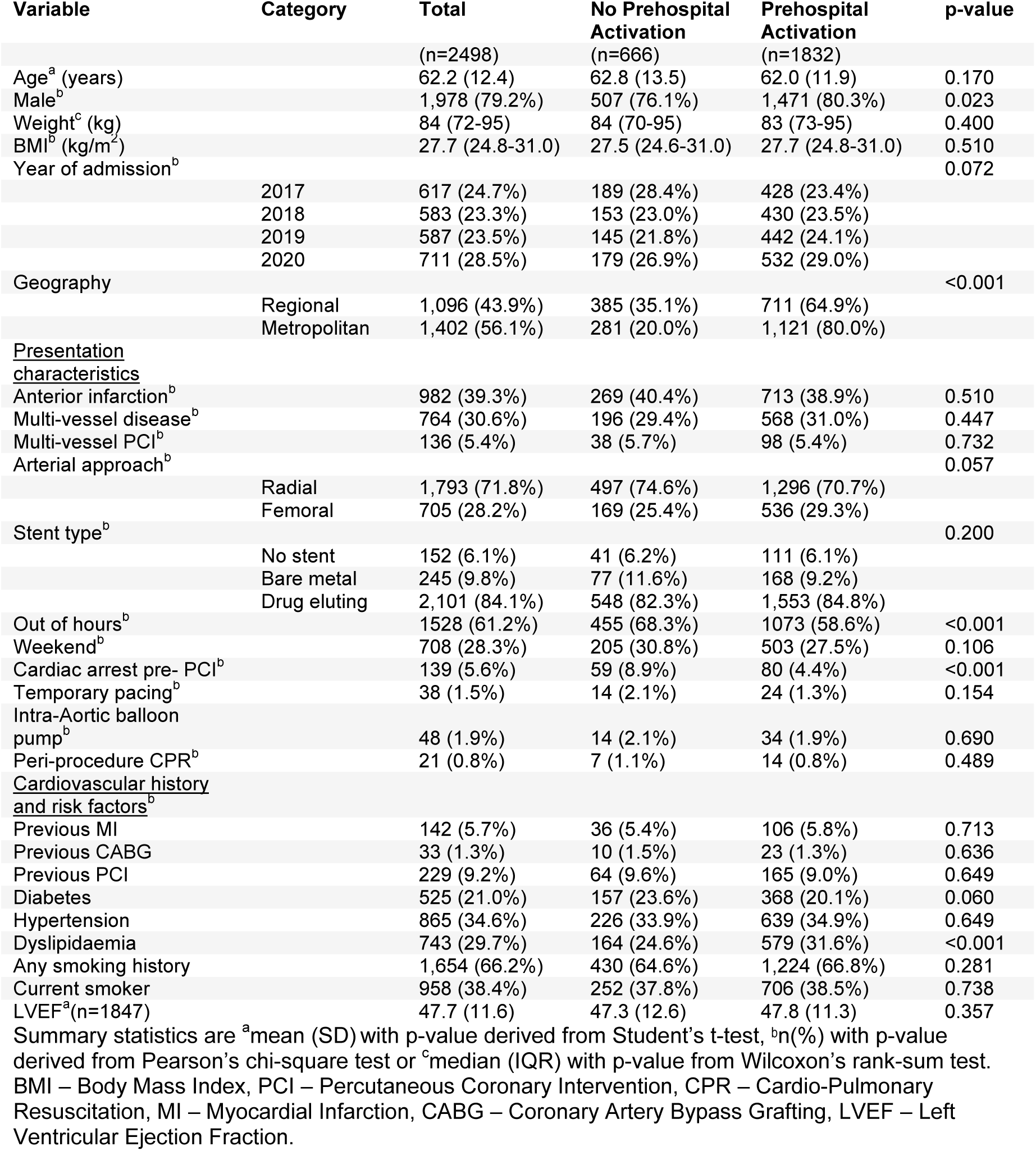
Summary statistics for patient characteristics by prehospital activation pathway.

### STEMI Performance Measures

Median DTB time was significantly shorter with prehospital activation (34mins vs 86mins; p<0.001) compared to those who did not have prehospital activation. Achievement of DTB<60mins (90% vs 16%; p<0.001) and FMC to balloon (43% vs 8%; p<0.001) and FDECG to balloon (62% vs 33%; p<0.001) targets were also both significantly improved with prehospital activation. Comparisons of total ischaemic time and critical timepoints between groups are displayed (Figure 2). There was a 48-minute difference in median hospital door to CCL time in prehospital activation patients compared to no prehospital activation (12mins vs 60mins; p<0.001). There was also a small yet significant difference in median CCL to balloon time (23mins vs 25mins; p<0.001) in the prehospital activation cohort. There was a 47-minute median reduction in FMC to balloon and 67-minute median reduction in in overall ischaemic time (Symptom to Balloon) with prehospital activation (Figure 2). Significant differences were seen in achievement of DTB<60mins both in hours (IH) and out of hours with prehospital activation associated with significant and comparable reductions in median DTB times (IH 74mins vs 29mins; p<0.001, OH 90mins vs 39mins; p<0.001) (Table 2).

**Figure 2.**
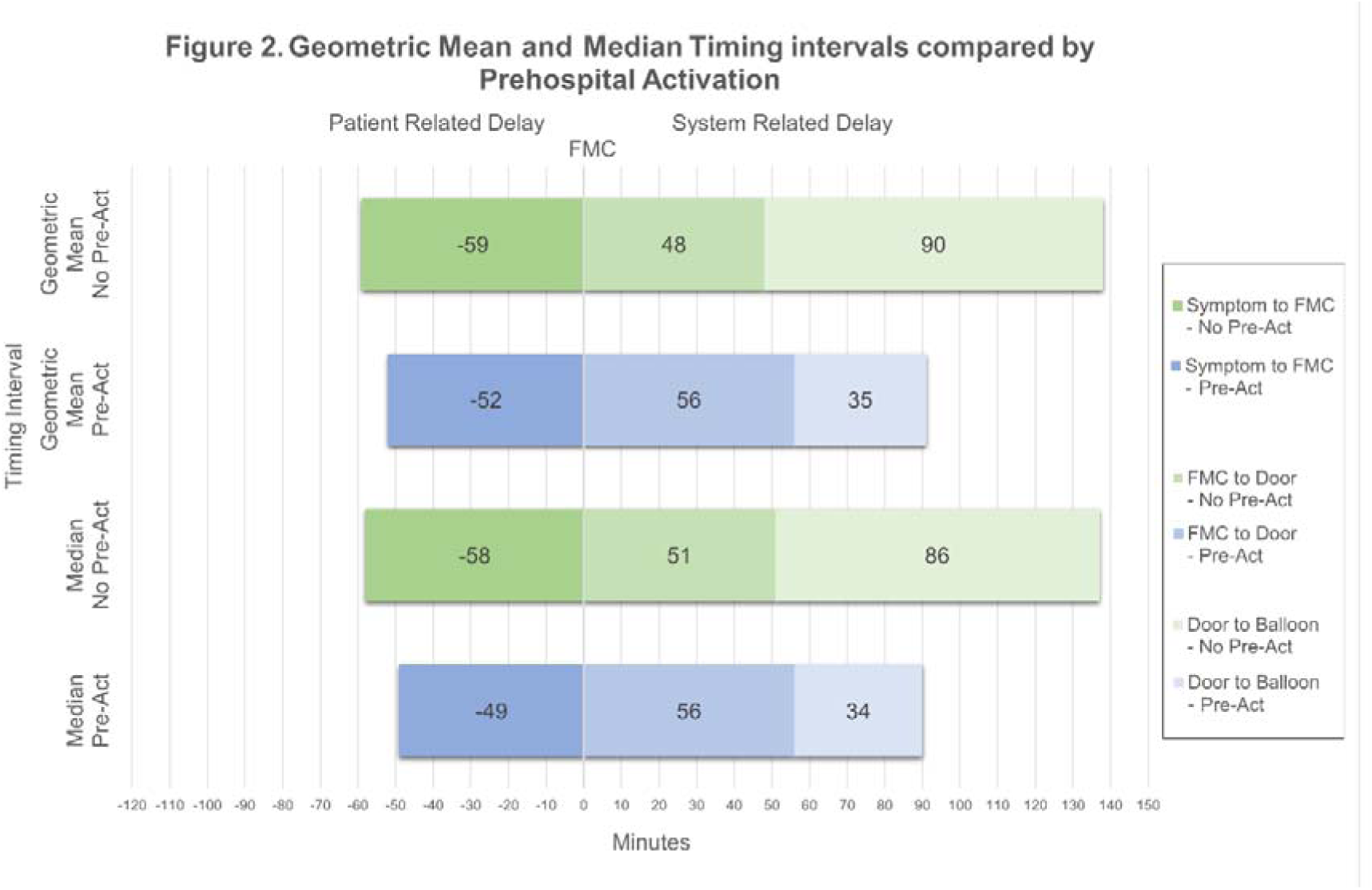
Comparison of geometric mean and median critical timepoints of ambulance transported STEMI patients undergoing primary PCI for STEMI. FMC – First Medical Contact. Pre-Act – Prehospital Activation. Prehospital activation cohort highlighted in blue. No prehospital activation cohort highlighted in green. Median and geometric mean times were similar across groups. Median time difference was 9 minutes shorter symptom onset to FMC, 5 minutes longer FMC to door time, and 52 minutes shorter DTB time with prehospital activation.

**Table 2:** Comparison of summary statistics^a^ for STEMI performance measures by prehospital activation pathway.

| Variable | Total | No Prehospital Activation | Prehospital Activation | p-value |
| --- | --- | --- | --- | --- |
|  | (n=2498) | (n=666) | (n=1832) |  |
| Symptom onset to FMC (minutes) | 51 (26-111) | 58 (29-126) | 49 (26-106) | 0.004 |
| FMC to FDECG (minutes) | 7 (4-15) | 13 (5-52) | 7 (4-11) | <0.001 |
| FMC to door (minutes) | 55 (45-67) | 51 (40-64) | 56 (47-67) | <0.001 |
| FMC to balloon (minutes) | 101 (84-127) | 140 (115-176) | 93 (80-109) | <0.001 |
| Symptom onset to balloon (minutes) | 165 (128-238) | 217 (168-295) | 150 (121-208) | <0.001 |
| Door to table (minutes) | 18 (9-40) | 60 (46-85) | 12 (7-22) | <0.001 |
| Table to balloon (minutes) | 24 (19-30) | 25 (20-31) | 23 (19-29) | <0.001 |
| DTB (minutes) | 42 (29-67) | 86 (68-113) | 34 (26-46) | <0.001 |
| FDECG to balloon (minutes) | 87 (73-108) | 109 (81-139) | 83 (72-98) | <0.001 |
| FMC to balloon<90Mins | 833 (33.3%) | 52 (7.8%) | 781 (42.6%) | <0.001 |
| FDECG to balloon<90Mins | 1,363 (54.6%) | 222 (33.3%) | 1,141 (62.3%) | <0.001 |
| DTB<60Mins | 1,752 (70.1%) | 107 (16.1%) | 1,645 (89.8%) | <0.001 |
| IH DTB (minutes) | 33 (25-52) | 90 (75-118) | 34 (26-46) | <0.001 |
| OH DTB (minutes) | 47 (33-77) | 74 (54-105) | 39 (30-50) | <0.001 |
| IH DTB<60Mins | 781 (80.5%) | 70 (33.2%) | 711 (93.7%) | <0.001 |
| OH DTB<60Mins | 971 (63.5%) | 37 (8.1%) | 934 (87.0%) | <0.001 |

<sup>a</sup>Summary statistics are median (IQR) with p-value from Wilcoxon's rank-sum test. FMC - First Medical Contact, FDECG - First Diagnostic Electrocardiograph, DTB - Door to Balloon.

### Mortality

Of 2498 patients, 74 (3.0%) died within 30-days (all cardiovascular cause) and 130 (5.2%) died within 1-year of the index procedure. Overall unadjusted 30-day mortality was significantly higher for patients who did not have prehospital activation compared to patients who had prehospital activation (6.6% vs 1.6%; p<0.001). Similarly, one-year all-cause mortality was significantly higher for those who did not have prehospital activation (10.2% vs 3.4%; p<0.001). One-year cardiovascular mortality was 4.2% and significantly higher for those who did not have prehospital activation (8.6% vs 2.6%; p<0.001). Based on summary statistics (Table 3), variables with strong unadjusted associations with 1-year mortality included age, diabetes, radial arterial access, drug-eluting stent usage, cardiac arrest pre-PCI, and FDECG to balloon time. After adjustment, patients who did not have prehospital activation were still at significantly increased risk of 30-day mortality compared to patients who had prehospital activation (total effect: OR: 3.6; 95%CI: 2.2-6.0, p<0.001; direct effect: OR: 2.9, 95%CI: 1.7-4.9; p<0.001), 1-year all-cause mortality (total effect: OR: 2.8; 95% CI: 1.9-4.1, p<0.001; direct effect: OR: 2.4; 95% CI: 1.6-3.6, p<0.001) and 1-year cardiovascular mortality (total effect: OR: 3.0; 95% CI: 2.1-5.1, p<0.001; direct effect: OR: 2.4; 95% CI: 1.5-3.8,p<0.001). Nelson-Aalen survival estimates by prehospital notification group are presented graphically in Figure 3. Consistent with the logistic regression results, there was markedly increased early mortality risk for those without prehospital activation compared to those with prehospital activation. After excluding the initial 30-day period, the difference in survival between groups amongst those who survived at least 30 days was not statistically significant (HR: 1.3; 95% CI: 0.9-2.1, P=0.21).

**Figure 3.**
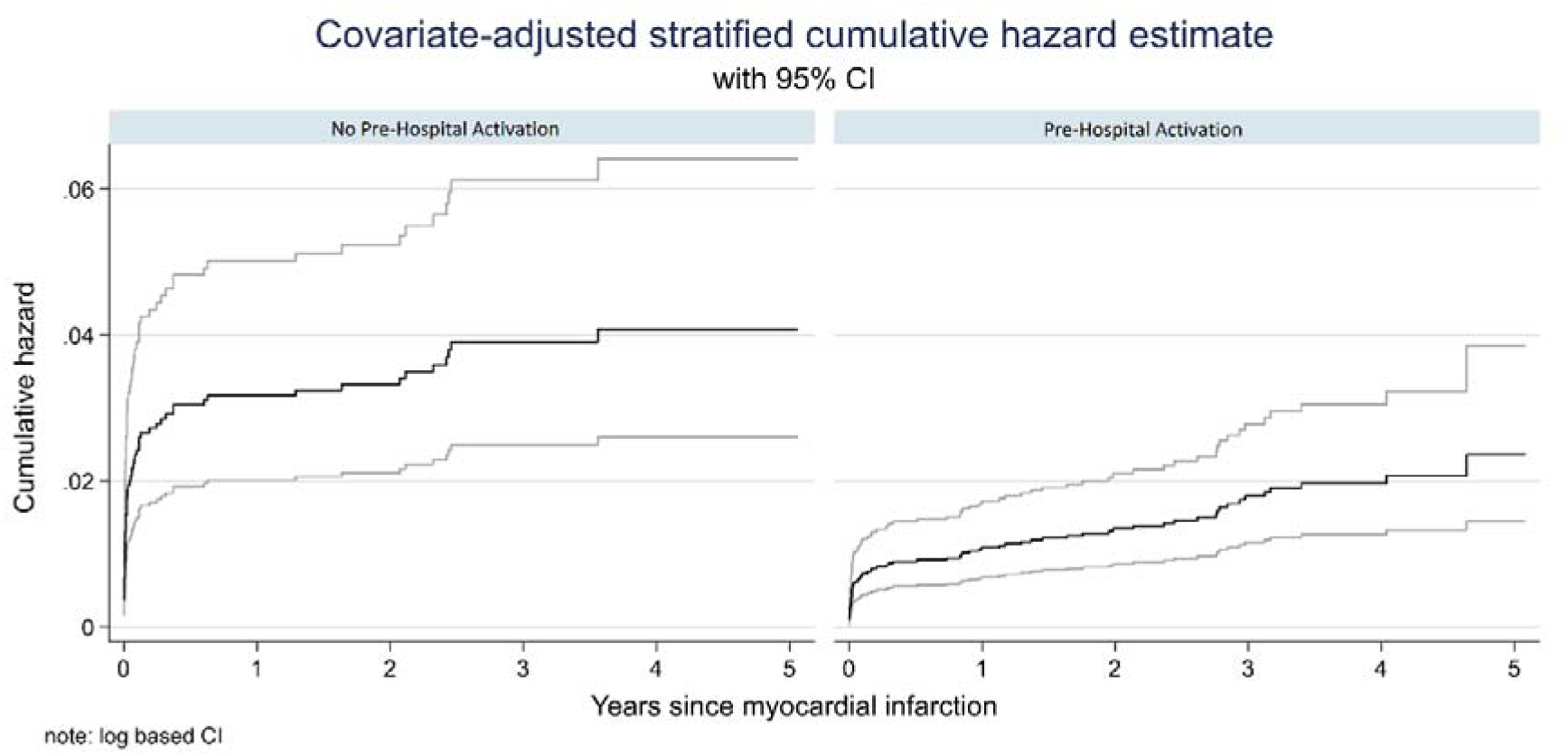
Adjusted cumulative hazard estimates for cardiovascular mortality with 95% confidence intervals by prior-hospital notification (PHN) status. Adjusted for age, sex, cardiac arrest pre-PCI, regional hospital and out of hours presentation.

**Table 3:** Summary statistics for variables of interest by 1-Year cardiovascular mortality status.

| Variable | Total Patients<br>(n=2498) | No<br>Cardiovascular<br>Death<br>(n=2369) | Cardiovascular<br>Death<br>(n=105) | p-value |
| --- | --- | --- | --- | --- |
| Age<sup>a</sup> (years) | 62.2 (12.4) | 61.8 (12.2) | 71.1 (13.8) | <0.001 |
| Male<sup>b</sup> | 1,978 (79.2%) | 1,900 (79.4%) | 78 (74.0%) | 0.208 |
| Weight<sup>c</sup> (kg) | 84.0 (72.0-95.0) | 84.0 (73.0-95.0) | 80.0 (67.5-90.5) | 0.013 |
| BMI<sup>c</sup> (kg/m<sup>2</sup>) | 27.7 (24.8-31.0) | 27.7 (24.8-31.0) | 26.6 (23.9-30.7) | 0.103 |
| Previous MI<sup>b</sup> | 142 (5.7%) | 137 (5.7%) | 5 (4.8%) | 0.677 |
| Previous PCI<sup>b</sup> | 229 (9.2%) | 214 (8.9%) | 15 (14.3%) | 0.063 |
| Previous CABG<sup>b</sup> | 33 (1.3%) | 32 (1.3%) | 1 (1.0%) | 0.735 |
| Diabetes<sup>b</sup> | 525 (21.0%) | 494 (20.6%) | 31 (29.5%) | 0.029 |
| Hypertension<sup>b</sup> | 865 (34.6%) | 822 (34.4%) | 43 (41.0%) | 0.164 |
| Dyslipidaemia<sup>b</sup> | 743 (29.7%) | 718 (30.0%) | 25 (23.8%) | 0.174 |
| Current smoker<sup>b</sup> | 958 (38.4%) | 926 (38.7%) | 31 (30.5%) | 0.090 |
| Radial access<sup>b</sup> | 1793 (71.8%) | 1729 (72.3%) | 64 (61.0%) | 0.012 |
| Drug eluting stent<sup>b</sup> | 2100 (84.1%) | 2023 (84.5%) | 77 (73.3%) | 0.002 |
| Anterior infarction<sup>b</sup> | 982 (39.3%) | 916 (38.3%) | 66 (62.9%) | <0.001 |
| Cardiac arrest pre PCI<sup>b</sup> | 139 (5.6%) | 123 (5.1%) | 16 (15.2%) | <0.001 |
| Multi-vessel disease<sup>b</sup> | 764 (30.6%) | 720 (30.1%) | 44 (41.9%) | 0.010 |
| LVEF%<sup>a</sup> (n=1847) | 48 (12) | 48 (11) | 36 (14) | <0.001 |
| Regional hospital<sup>b</sup> | 1,096 (43.9%) | 1,046 (43.7%) | 50 (47.6%) | 0.624 |
| Out of hours<sup>b</sup> | 1,528 (61.2%) | 1,458 (60.9%) | 70 (66.7%) | 0.238 |
| Weekend presentation<sup>b</sup> | 708 (28.3%) | 684 (28.6%) | 24 (22.9%) | 0.203 |
| Prehospital activation<sup>b</sup> | 1832 (73.3%) | 1784 (74.6%) | 48 (45.7%) | <0.001 |
| DTB<sup>c</sup> (mins) | 42 (29-67) | 41 (29-65) | 68 (38-94) | <0.001 |
| FMC to balloon<sup>c</sup> (mins) | 101 (84-127) | 100 (84-125) | 128 (99-160) | <0.001 |
| FDECG to balloon<sup>c</sup> (mins) | 87 (73-108) | 86 (73-106) | 108 (87-140) | <0.001 |
| STB<sup>c</sup> (mins) | 165 (128-238) | 163 (127-237) | 191 (146-260) | 0.005 |

Summary statistics are <sup>a</sup> mean (SD) with p-value derived from Student's t-test, <sup>b</sup> n(%) with p-value derived from Pearson's chi-square test, or <sup>c</sup> median (IQR) with p-value from Wilcoxon's rank-sum test. BMI – Body Mass Index, MI – Myocardial Infarction, PCI – Percutaneous Coronary Intervention, CABG – Coronary Artery Bypass Graft, LVEF – Left Ventricular Ejection Fraction, DTB – Door to Balloon, FMC – First Medical Contact, FDECG – First Diagnostic Electrocardiograph, STB – Symptom to Balloon.

**Table 4:** Mortality outcomes and logistic regression modelling for the effects of prehospital activation on 1-year cardiovascular mortality.

| Outcome Variable | Total<br>(n=2498) | No Prehospital<br>Activation<br>(n=666) | Prehospital<br>Activation<br>(n=1832) | p-value |
| --- | --- | --- | --- | --- |
| 30 Day All-Cause Mortality<sup>a</sup> | 74 (3.0%) | 44 (6.6%) | 30 (1.6%) | <0.001 |
| 1 Year All-Cause Mortality<sup>a</sup> | 130 (5.2%) | 68 (10.2%) | 62 (3.4%) | <0.001 |
| 1 Year CV Mortality<sup>a</sup> | 105 (4.2%) | 57 (8.6%) | 48 (2.6%) | <0.001 |

| Exposure Variable | Category | Unadjusted effect<br>OR (95 CI) | p-value | Direct effect<sup>b</sup><br>OR (95 CI) | p-value | Total effect<sup>b</sup><br>OR (95 CI) | p-value |
| --- | --- | --- | --- | --- | --- | --- | --- |
| Prehospital activation | Yes | Reference |  | Reference |  | Reference |  |
|  | No | 3.5 (2.3-5.2) | <0.001 | 2.3 (1.5-3.7) | <0.001 | 3.0 (2.0-4.6) | <0.001 |
| Age (years) |  |  | <0.001 |  | <0.001 |  | <0.001 |
|  | <66 | Reference |  | Reference |  | Reference |  |
|  | 65-<75 | 1.1 (0.7-2.0) | 0.738 | 1.1 (0.6-2.0) | 0.721 | 1.2 (0.7-2.1) | 0.569 |
|  | ≥75 | 5.3 (3.4-8.3) | <0.001 | 5.3 (3.3-8.4) | <0.001 | 5.7 (3.6-9.1) | <0.001 |
| Cardiac arrest pre-PCI | No | Reference |  | Reference |  | Reference |  |
|  | Yes | 3.3 (1.9-5.8) | <0.001 | 3.2 (1.8-5.9) | <0.001 | 3.5 (1.9-6.3) | <0.001 |
| Out of hours presentation | Yes | Reference |  | Reference |  | Reference |  |
|  | No | 0.8 (0.5-1.2) | 0.239 | 0.9 (0.6-1.4) | 0.602 | 0.8 (0.5-1.3) | 0.335 |
| Sex | Female | Reference |  | Reference |  | Reference |  |
|  | Male | 0.8 (0.5-1.2) | 0.208 | 1.1 (0.7-1.8) | 0.703 | 1.1 (0.7-1.7) | 0.777 |
| Regional hospital | No | Reference |  | Reference |  | Reference |  |
|  | Yes | 1.2 (0.8-1.7) | 0.430 | 0.9 (0.6-1.4) | 0.731 | 0.8 (0.6-1.3) | 0.388 |
| FDECG to balloon (mins) |  |  | <0.001 |  | 0.016 |  |  |
|  | <90 | Reference |  | Reference |  |  |  |
|  | 90-<120 | 1.5 (0.9-2.5) | 0.130 | 1.2 (0.7-2.0) | 0.559 |  |  |
|  | ≥120 | 4.3 (2.8-6.9) | <0.001 | 2.1 (1.2-3.7) | 0.006 |  |  |

<sup>a</sup>Summary statistics are n (%) with p-value derived from Pearson's chi-square test. CV – Cardiovascular, PCI – Percutaneous coronary intervention, FDECG – First Diagnostic Electrocardiograph. <sup>b</sup>Note that inference should be limited to the exposure of interest; it is not appropriate to interpret the effects of other covariates in these multivariable models.

## Discussion

This state-wide multicentre registry study highlights the low mortality rates and significant improvements in STEMI performance measures associated with patients who underwent ambulance prehospital activation and direct catheterisation laboratory transfer for STEMI. Previously published literature regarding the impact of prehospital activation of STEMI patients are consistent with the findings of this study with the consensus that there are significantly improved STEMI performance metrics [8, 9, 11, 12, 15, 22, 23]. Whilst the literature is predominantly supportive of lower mortality associated with prehospital activation strategies [8, 12–15], recently published data was suggestive of no mortality benefit with a prehospital notification strategy [16, 17]. The published literature from Australia regarding mortality is mixed with national data showing lower mortality with ambulance transported patients compared to those who self-present in the GRACE and CONCORDANCE registries [24]. More recent data from Victoria however highlighted lower mortality with patients who self-presented compared to ambulance transported patients, however there were significantly greater comorbidities, higher rates of cardiogenic shock and mechanical ventricular support in the ambulance transported groups in this cohort study [17]. The increased risk of mortality with associated no prehospital activation in our study are consistent with mortality estimates demonstrated in previous literature with increasing risk of mortality associated with delays to treatment [25, 26].

Prehospital anti-thrombotic medication has been associated with improved TIMI flow prior to PCI, lower rates of 30-day stent thrombosis and lower mortality [27–29] and may explain, in-conjunction with lower time to reperfusion, the association with lower mortality with prehospital activation in our study compared to previously published Australian literature [16, 17]. While these studies did demonstrate similar reductions in time to reperfusion, it is unclear the timing of initiation of medical therapy, including anti-thrombotic medication which may also contribute to the mortality differences. Other differences in mortality in previously reported studies may be due to reporting all-cause mortality rather than specifically cardiovascular mortality as well as the inclusion/exclusion of patients with cardiogenic shock, out-of-hospital cardiac arrest and pre-procedural intubation with uncertain neurological status. Our study excluded patients who had out-of-hospital cardiac arrest <u>and</u> pre-procedural intubation with uncertain neurological status. These patients were deemed salvage due to the significantly high proportion of neurovascular mortality in this group despite appropriate cardiovascular reperfusion. However, the current study did not exclude patients with cardiogenic shock which may develop with longer delays to treatment as noted in other studies [17]. Cardiac arrest prior to PCI was identified on the causal pathway and was incorporated into the multivariate analysis. Unlike previous studies, cause of death was also analysed in our study and both all-cause and cardiovascular mortality showed lower rates associated with prehospital activation.

Significant reductions in all STEMI performance measures were associated with prehospital activation. Whilst the current Australian [18] and ACC/AHA STEMI [30] guidelines suggest the metric FMC to balloon, the ESC STEMI guidelines [1] have adopted a STEMI diagnosis to device time within 90 minutes as a performance measure. This study also incorporated the similar metric of FDECG to balloon time, as well as FMC to balloon, which may be a more achievable performance metric when compared to FMC to balloon especially when there is delayed electrocardiographic or evolving evidence of STEMI. There was a significant albeit small difference in our study in median time from FMC to FDECG between groups with noticeably shorter time in the prehospital activation cohort. This may be explained in part by evolving or delayed electrocardiographic evidence of STEMI, however it is important to acknowledge that initial misdiagnosis of STEMI may contribute to this, despite prehospital activation being associated with lower false positive STEMI activations [9, 10, 14]. Prehospital activation had the largest system timing impact on DTB time (Figure 2). Whilst the focus has shifted to overall system delays, DTB time still provides actionable targets to hospitals and is directly affected by in-hospital processes. This metric can still be used to support in-hospital improvements and accountability [31]. Similar to previously published data there was a 5-minute difference in the median FMC to Door time with patients who did not have prehospital activation being slightly faster (Figure 2). Additional workload performed by paramedics in the prehospital activation pathway (patient consent, primary PCI referral with the interventional cardiologist and subsequent administering of medication), may account for these differences.

The proportion of patients with STEMI who are ambulance transported and have prehospital notification/activation in published Australian literature continues to remain low and constant with less than half of patients receiving prehospital notification [9, 16, 24, 32]. This is significantly lower than current data presented in this study (73%). Recent international data from the United States is also suggestive of lower rates of prehospital activation compared to the current study with approximately 40% of patients having prehospital activation [7]. A comparison of prehospital activation strategies between Queensland and the Mission Lifeline: Pre-ACT strategy is displayed in Figure 4. There were regional differences in the use of prehospital activation in our study which may be associated with numerous factors such as paramedic staffing and experience levels as well as other unobserved factors. Increasing the use of prehospital activation may present as an opportunity to improve STEMI care in regional areas. Previous examination of predictors of prehospital notification and similar to our study demonstrated greater use of prehospital notification during in-hours [17], yet unlike the predictors listed in this Victorian study, our study did not associate any sex-related differences with prehospital activation nor differences in the identified culprit coronary vessel. Previous literature examining sex differences in STEMI have suggested the use of standardised prehospital protocols reducing the differences in sex related treatment discrepancies [33, 34].

**Figure 4.**
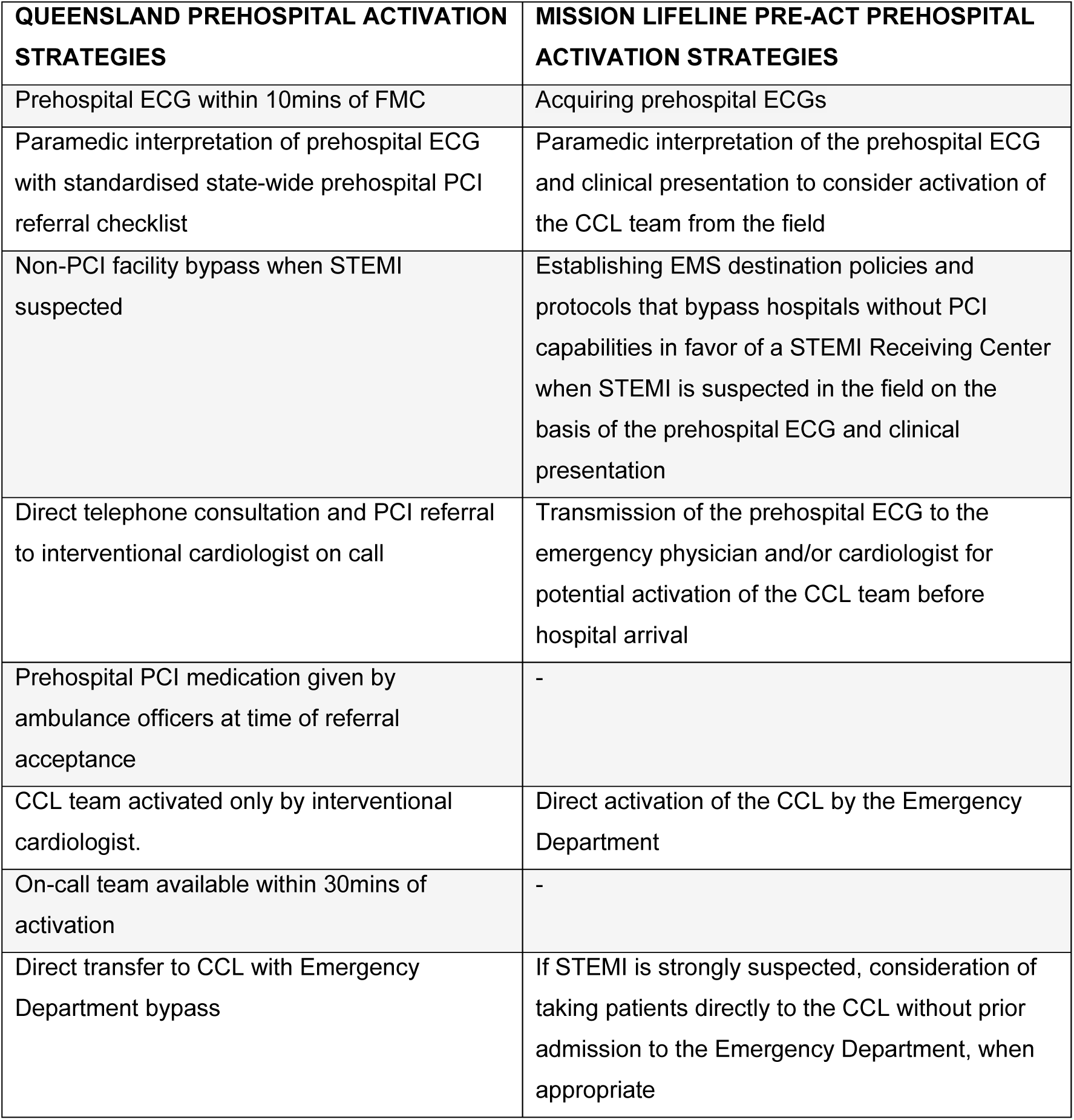
Comparison of prehospital activation strategies between the Queensland and Mission Lifeline PRE-ACT strategies. FMC – First Medical Contact, ECG – Electrocardiograph, PCI – Percutaneous Coronary Intervention, STEMI – ST-segment Elevation Myocardial Infarction, CCL – Cardiac catheterisation laboratory.

This study adds to the support for standardised prehospital strategies incorporating prehospital notification of STEMI, initiation of medical therapy and activation of the cardiac catheterisation laboratory to improve STEMI performance measures, reduce overall time to reperfusion and lower both short- and longer-term mortality. Routine monitoring of prehospital activation rates may provide opportunities to improve prehospital care in STEMI.

## Strengths and Limitations

This is a multicentre study analysing prospectively collected data from the QCOR registry in a large cohort of consecutive STEMI patients treated with Primary PCI. Despite our statistical methods used and due to the observational nature of this study, there may be unmeasured cofounders which could influence mortality. Whilst there may be unmeasured confounders, our study did have complete and high-quality measures for many important confounders related to cardiovascular outcomes as described in the causal pathway diagram. A comparison of total and direct effects involved in the association between prehospital activation and 1-year mortality enables us to better understand the causal pathways.

Selection bias may exist when identifying STEMI in the prehospital setting and may result in exclusion of ambiguous or evolving STEMI presentations or those patients where STEMI was not initially recognised, from the direct primary PCI referral pathway. Previously documented reasons for no prehospital activation include availability of critical care paramedics [14] which may have contributed to the lower utilization of prehospital activation in more regional centres and after-hours as well as proximity to hospital, which also may explain the significantly shorter FMC to hospital arrival time in the no-prehospital activation group [35]. Additionally bypass of non-PCI facilities could contribute to longer prehospital travel times in the prehospital notification group and has been previously demonstrated to increase ambulance transport time [36]. Our study also only compared patients who underwent primary PCI for treatment of STEMI within 12 hours of symptom onset and STEMI patients who were treated medically or with coronary artery bypass may add to potential bias. This study did not analyse secondary prevention measures such as post procedural medication compliance or cardiac rehabilitation adherence which may also influence longer-term mortality.

## Conclusion

A standardised state-wide strategy incorporating prehospital notification of STEMI, initiation of medical therapy and prehospital activation of the cardiac catheter laboratory significantly improved STEMI performance measures, reduced total ischaemic time and is associated with lower 30-day and 1-year mortality. Widespread implementation of prehospital activation may offer significant opportunity to expedite STEMI care and improve outcomes.

## Data Availability

The clinical data used to support the findings of this study are restricted by the Prince Charles Hospital Human research and ethics committee in order to protect patient privacy and confidentiality. Data are available from the relevant Prince Charles data custodian/s for researchers who meet the criteria for access to confidential data.

## Acknowledgements

Nil

## Ethics Approval/s

This study was approved by the Human Research Ethics Committee of the Prince Charles Hospital (LNR/2020/TPCH/75136).

---

**Supplement 1.**
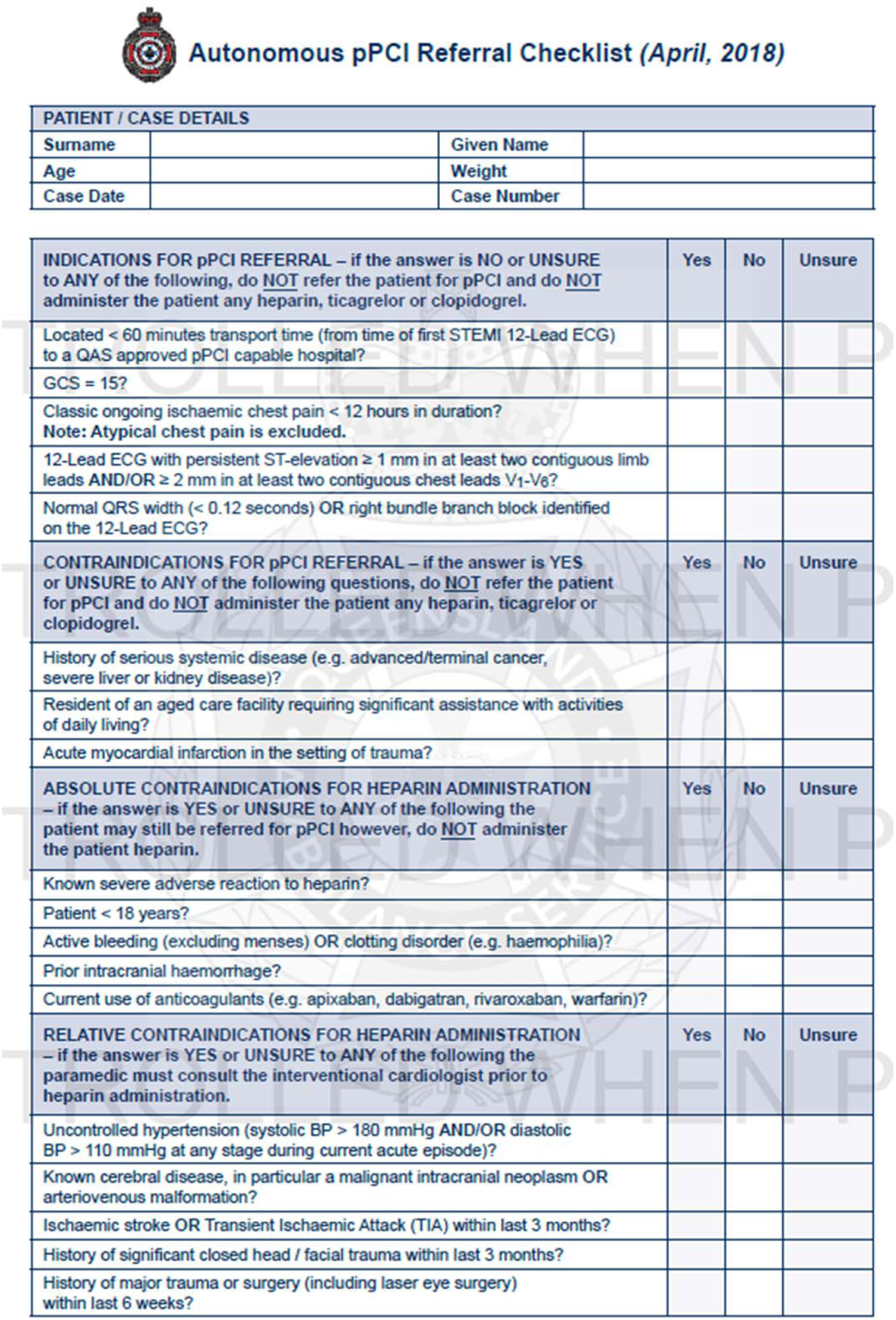
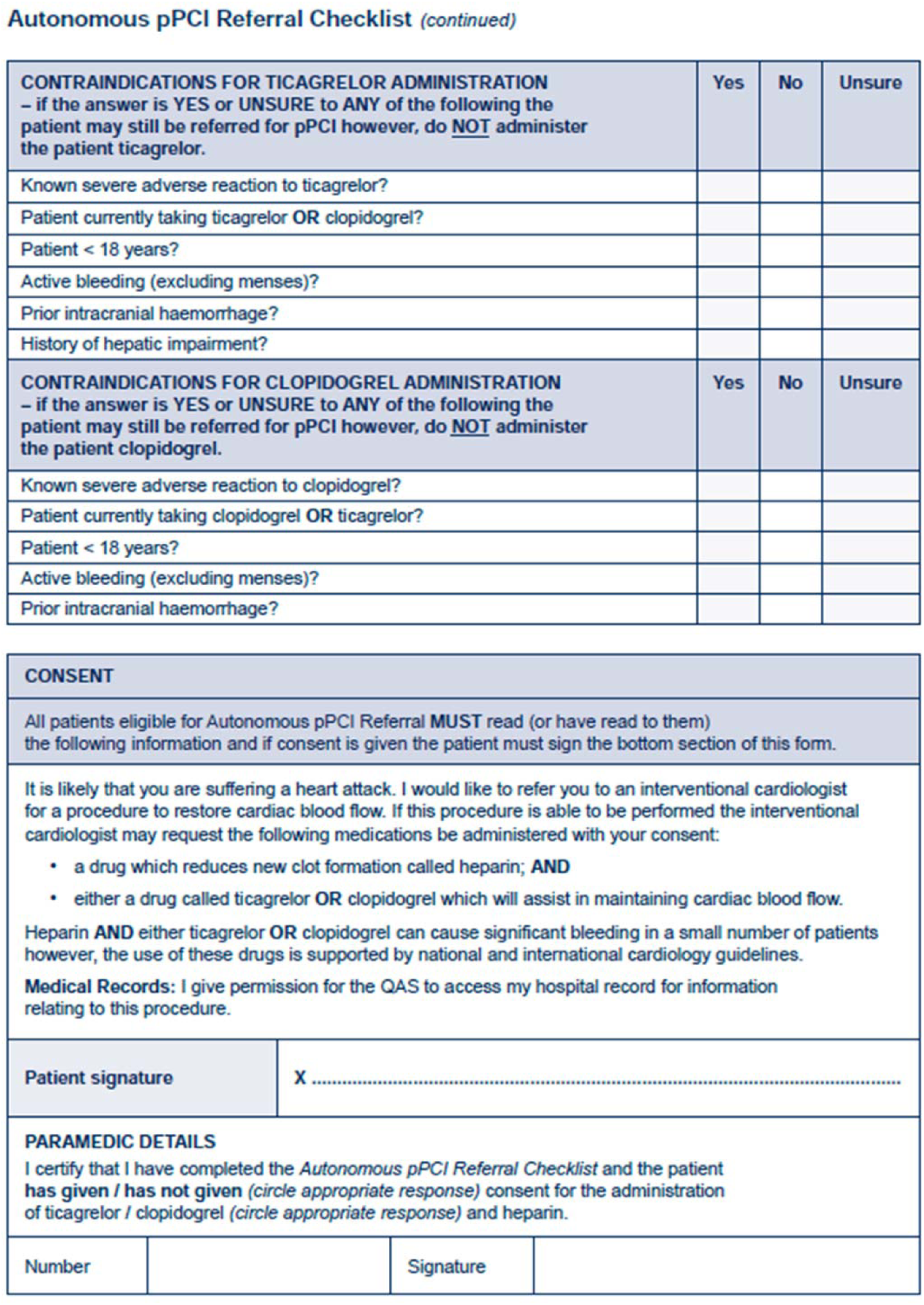
Queensland Ambulance Service – Autonomous primary PCI referral Checklist.

**Supplement 2.**
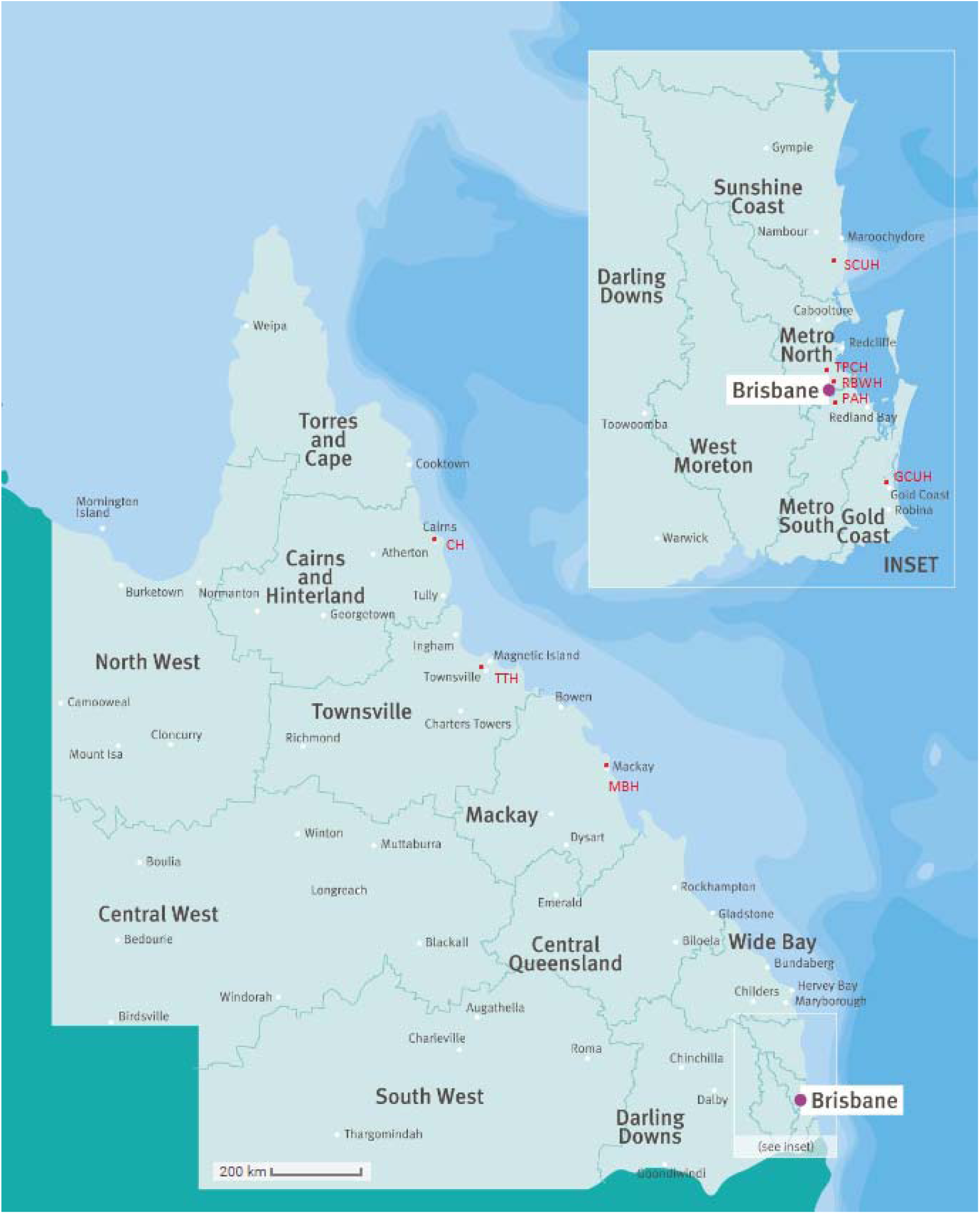
Queensland Map demonstrating participating primary PCI centres. Queensland map demonstrating Metropolitan centres (TPCH-The Prince Charles Hospital, RBWH – Royal Brisbane and Women’s Hospital, PAH – Princess Alexandra Hospital) and Regional centres (CH – Cairns Hospital, TTH – Townsville Hospital, MBH – Mackay Base Hospital, SCUH – Sunshine Coast University Hospital, GCUH – Gold Coast University Hospital)

**Supplement 3.**
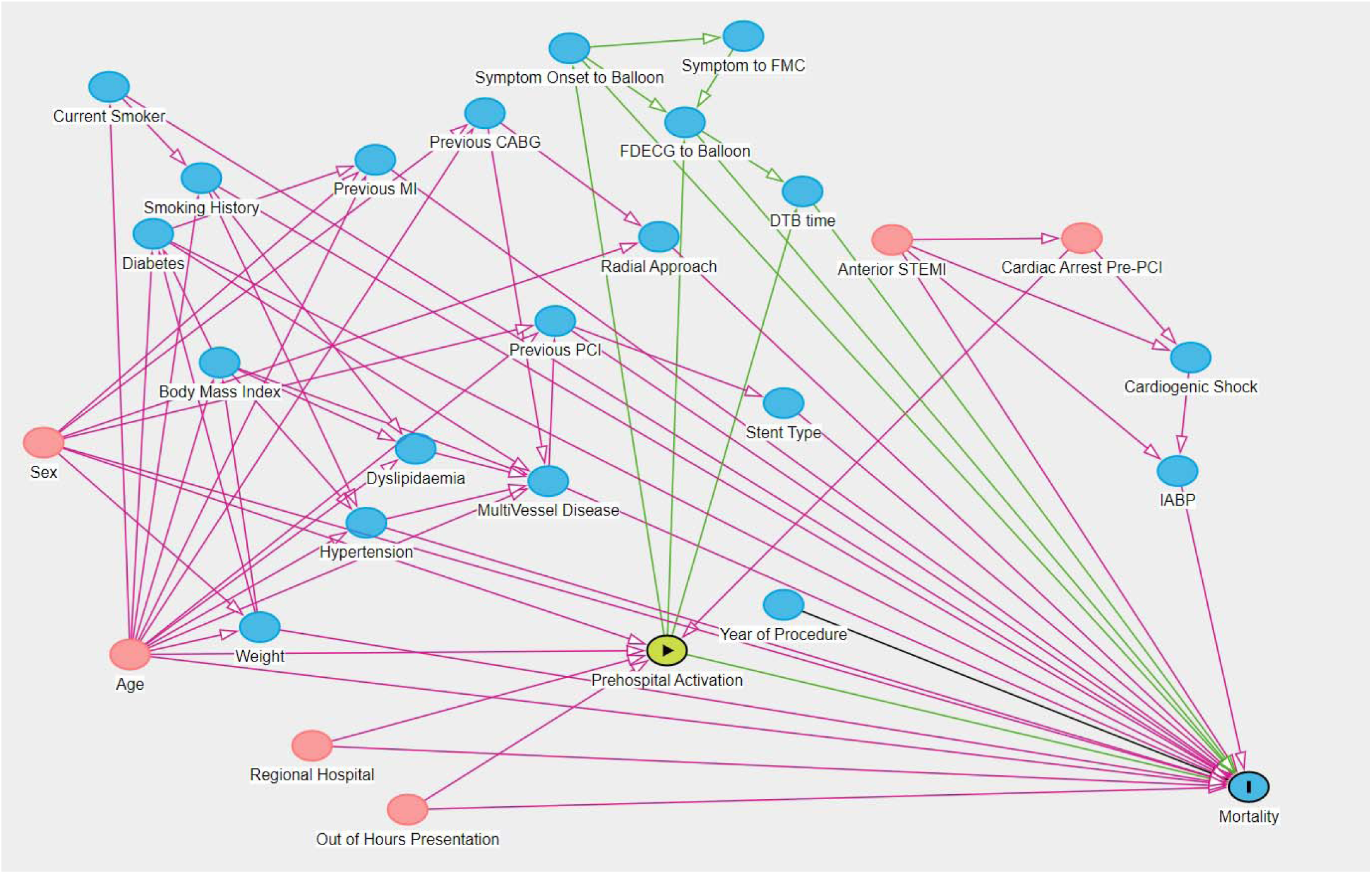
Dagitty causal pathway diagram. MI – Myocardial Infarction, PCI – Percutaneous Coronary Intervention, CABG – Coronary Artery Bypass Graft, DTB – Door to Balloon, FMC – First Medical Contact, FDECG – First Diagnostic Electrocardiograph, STB – Symptom to Balloon, STEMI – ST-segment Elevation Myocardial Infarction, IABP – Intra-aortic Balloon Pump.

